# Whole Genome Sequencing for the Diagnosis of Rare Disorders

**DOI:** 10.1101/2025.04.25.25326373

**Authors:** Antonio Victor Campos Coelho, Rafael Sales de Albuquerque, Catarina dos Santos Gomes, José Bandeira do Nascimento Junior, Gustavo Santos de Oliveira, Livia Maria Silva Moura, Luciana Souto Mofatto, Rafael Lucas Muniz Guedes, Rodrigo Araújo Sequeira Barreiro, Marcel Pinheiro Caraciolo, Ana Paula de Andrade Oliveira, Anne Caroline Barbosa Teixeira, Bruna Mascaro Cordeiro de Azevedo, Carolina Dias Carlos, Lucas Santos de Santana, Marina Cadena da Matta, Matheus Martinelli Lima, Nuria Bengala Zurro, Renata Yoshiko Yamada, Vivian Pedigone Cintra, Gabriela Pereira Campilongo, Gabriela Borges Cherulli Colichio, Renata Martins Ribeiro da Silva, Caio Robledo D’Angioli Costa Quaio, Carolina Araújo Moreno, Eduardo Perrone, Jéssica Grasiela Araújo Espolaor, Joana Rosa Marques Prota, José Ricardo Magliocco Ceroni, Kelin Chen, Luiza do Amaral Virmond, Marina de Franca Basto Silva, Michele Patricia Migliavacca, Renata Moldenhauer Minillo, Thiago Yoshinaga Tonholo Silva, Karla de Oliveira Pelegrino, Ana Luiza Garcia Cunha, Joziele de Souza Lima, Anete Sevciovic Grumach, Caio Parente Barbosa, Angelina Xavier Acosta, Paula Brito Corrêa, Denise Pontes Cavalcanti, Carlos Eduardo Steiner, Erlane Marques Ribeiro, Wallace William da Silva Meireles, Giselle Maria Araujo Felix Adjuto, Ida Vanessa Doederlein Schwartz, Têmis Maria Felix, Irma Cecilia Douglas Paes Barreto, Antonette Souto El-Husny, Jussara Melo de Cerqueira Maia, Vera Maria Dantas, Lúcia Helena de Oliveira Cordeiro, Luisa Zagne Braz, Magda Maria Sales Carneiro Sampaio, Mara Lucia Schmitz Ferreira Santos, Marco Antonio Curiati, Maria Teresinha de Oliveira Cardoso, Maria Teresa Alves da Silva Rosa, Mariana Paes Leme Ferriani, Ester Silveira Ramos, Paula Teixeira Lyra, Raquel Tavares Boy da Silva, Anna Cândida Ximenes de Mendonça Sobreira, Tatiana Regia Suzana Amorim Boa Sorte, Melissa Rossi Calvão Dumas, Thaís Bomfim Teixeira, Vandré Cabral Gomes Carneiro, Patrícia Silva Mota, Tatiana Ferreira de Almeida, João Bosco Oliveira

**Affiliations:** Hospital Israelita Albert Einstein, Avenida Albert Einstein 627, São Paulo 05652-900, Brazil; Universidade Federal de São Paulo, Departamento de Morfologia e Genética, Rua Botucatu 740, São Paulo 04023-000, Brazil; Hospital Infantil João Paulo II - FHEMIG, Alameda Ezequiel Dias 345, Belo Horizonte 30130-110, Brazil; Centro Universitário Faculdade de Medicina do ABC, Avenida Lauro Gomes 2000, Santo André 09060-870, Brazil; Hospital Prof. Edgar Santos (Hupes-UFBA), Avenida Augusto Viana Setor Genética Médica s/n, Salvador 40110-060, Brazil; Departamento de Genética Médica, FCM, Universidade Estadual de Campinas (UNICAMP), Rua Tessália Vieira de Camargo 126, Campinas 13083-887, Brazil; Hospital Infantil Albert Sabin (HIAS), Rua Tertuliano Sales 544, Fortaleza 60410-794, Brazil; Hospital de Apoio de Brasília (HAB DF), Setor Noroeste AENW Lote A 3, Brasília 70684-831, Brazil; Hospital de Clínicas de Porto Alegre (HCPA), Rua Ramiro Barcelos 2350, Porto Alegre 90035-903, Brazil; Centro Universitário do Estado do Pará (CESUPA), Avenida Governador José Malcher 1242, Belém 66060-230, Brazil; Hospital Universitário Onofre Lopes (HUOL/UFRN), Avenida Nilo Peçanha 620, Natal 59012-300, Brazil; Departamento de Medicina Clínica, Universidade Federal de Pernambuco (UFPE), Avenida Professor Moraes Rego 1235, Recife 50670-901, Brazil; Hospital das Clínicas da Faculdade de Medicina da Universidade de São Paulo (HCFMUSP), Rua Doutor Ovídio Pires de Campos 225, São Paulo 05403-010, Brazil; Hospital de Crianças César Pernetta e Hospital Pequeno Príncipe, Rua Desembargador Motta 1070, Curitiba 80250-060, Brazil; Universidade Federal de São Paulo, Centro de Referência em Erro Inato do metabolismo (CREIM), Rua coronel Lisboa 957, São Paulo 04020-040, Brazil; Centro de Referência em Doenças Raras (UGEN HAB/SES-DF), Avenida L2 Sul SGAS Quadra 608 - Módulo A, Brasília 70203-900, Brazil; Universidade Católica De Brasília (UCB), QS 07 Lote 01, Brasília 71966-700, Brazil; Hospital das Clínicas da Faculdade de Medicina de Ribeirão Preto (HCFMRP), Rua Tenente Catão Roxo 3900, Ribeirão Preto 14015-010, Brazil; Instituto de Medicina Integral Professor Fernando Figueira (IMIP), Rua dos Coelhos 300, Recife 50070-902, Brazil; Hospital Universitário Pedro Ernesto - Universidade do Estado do Rio de Janeiro (HUPE/UERJ), Boulevard Vinte e Oito de Setembro 77, Rio de Janeiro 20551-030, Brazil; Associação de Pais e Amigos dos Excepcionais - Salvador (APAE Salvador), Rua Espírito Santo 575, Salvador 41830-120, Brazil; Associação de Pais e Amigos dos Excepcionais - Anápolis (APAE Anápolis), Rua Galileu Batista Arantes 350, Anápolis 75075-570, Brazil; Hospital de Câncer de Pernambuco (HCPE), Avenida Cruz Cabugá 1597, Recife 50040-000, Brazil; Previously Hospital Israelita Albert Einstein, currently Neogenomica Análises Genômicas, Rua Frei Matias Teves 285, Recife 50070-465, Brazil

## Abstract

**Background:** Whole-Genome Sequencing (WGS) and Whole-Transcriptome Sequencing (WTS) have emerged as transformative tools in the diagnosis of rare diseases with complex phenotypes. These technologies enable deep analysis of the genome and RNA expression, uncovering structural, intronic, non-coding, and mitochondrial variants that traditional methods might miss, thus facilitating the understanding of gene function and regulation.

**Methods:** We enrolled 8966 patients with suspected rare diseases or hereditary cancer risk syndromes from 21 centers throughout Brazil. Their genomes were sequenced with short, paired-end reads, and diagnostic reports were provided for 7614 of these patients.

**Results:** The overall diagnostic yield was 35.8%, with an additional 5.6% of all Positive reports including gains from WGS compared to other diagnostic tests. In patients with pathogenic/likely pathogenic copy number or structural variants, WGS provided a 10.2% increase in diagnostic yield. Almost 1900 variant/phenotype interpretations were submitted to ClinVar.

**Conclusion:** WGS and WTS are proving to be invaluable resources for shortening the diagnostic odyssey of patients with rare diseases, providing crucial genomic diagnostics, and enriching genetic databases with variant interpretations from underrepresented populations. These technologies have the potential to significantly enhance the precision of healthcare in genetically diverse populations.

**Key Points for NEJM Editors and Reviewers:**

1. **Scope**: Largest Brazilian rare disease–focused sequencing project to date.
2. **Findings**: A 35.8% diagnostic yield with notable additional detection of structural, deep intronic, non-coding, and mitochondrial variants.
3. **Impact**: Shortens diagnostic odyssey for patients, enriches ClinVar with almost 1900 variant interpretations from underrepresented populations, and demonstrates potential cost-effectiveness of integrating WGS into public healthcare. These interim results underscore how national WGS programs can facilitate precision medicine, especially in genetically diverse and previously underrepresented populations.

## 1. Introduction

Rare diseases have an estimated prevalence ranging between 1.5% and 6.2% in the general population ^1^ which translates to between 3.2 and 13.2 million affected individuals in Brazil alone. Brazil, notable for its free-of-charge health care system (the *Sistema Único de Saúde*, SUS) ^2^, has made significant strides in improving rare disease diagnosis since 2014. Brazilian healthcare centers now offer newborn metabolic screening and genetic tests such as karyotyping, FISH, MLPA, and CGH-array.

However, whole-genome sequencing (WGS) and whole-transcriptome sequencing (WTS) have emerged as transformative tools in the diagnosis of rare diseases with complex phenotypes. WGS enables deep analysis of the genome, uncovering structural, intronic, non-coding, and mitochondrial variants that traditional methods might miss. Similarly, WTS provides a comprehensive view of RNA expression, facilitating the understanding of gene function and regulation.

Countries like the United Kingdom, Australia, Belgium, Canada, France, and many others have pioneered genetics-based precision medicine national programs ^3–5^, underscoring the global recognition of these technologies’ diagnostic potential. Unfortunately, Brazilian populations have been underrepresented in these international genomics initiatives.

This study investigates the diagnostic power of WGS and WTS within the Brazilian public healthcare system in the context of the Rare Genomes Project (BRGP), demonstrating their potential to significantly shorten the diagnostic odyssey for patients and enrich genetic databases with variant interpretations from underrepresented populations. The findings presented herein highlight the high value of integrating these advanced genomic tools into public healthcare.

## 2. Methods

The overall inclusion criteria include a suspicion of rare disease with probable genetic etiology or hereditary cancer risk syndromes.

For pediatric patients, parents or legal guardians are asked for consent before inclusion, with additional consent required for secondary findings investigation. Patients are excluded from the project if they refuse the invitation for inclusion, have a prior positive molecular or karyotypic diagnosis, or if the condition is likely caused by environmental factors.

Once patients or their families provide written informed consent, their information is stored electronically in the REDCap ^6,7^ and PhenoTips ^8^ databases. Physicians then assign each patient to one of 20 disorder cohorts based on specific criteria (Neurologic disorders, Neuromuscular disorders, Neurocutaneous disorders, Inborn errors of metabolism, Skeletal disorders, Hereditary cancer risk syndromes, Clinically recognizable genetic syndromes, Endocrine disorders, Connective tissue disorders, Immunologic disorders, Ophthalmologic disorders, Hearing or ear disorders, Renal and urinary tract disorders, Dermatologic disorders, Hematologic disorders, Cardiological disorders, Pulmonary disorders, Gastroenterologic disorders, Vascular disorders, and Critically ill patients – children aged one year or younger admitted to neonatal or pediatric intensive care units). They also provide a detailed clinical summary, including indications for referral, pedigrees, therapeutic approach, and Human Phenotype Ontology (HPO) terms ^9^.

Following enrollment, patients provide blood samples for genomic DNA automated extraction. The sequencing protocol is detailed elsewhere ^10^. Briefly, PCR-free WGS is performed on Illumina NovaSeq 6000 equipment. Read mapping and variant call ared done using the GRCh38 build reference genome. The VCF files are made available for analysis on the Varstation® platform (Genesis Genomics, São Paulo, Brazil) ^11^, which annotates the VCF files using Ensembl’s Variant Effect Predictor (VEP) ^12^ and AnnotSV ^13,14^. Additionally, for a limited time during the second triennial phase of the BRGP, we obtained a single PAXgene® Blood RNA tube vial (BD Biosciences, Franklin Lakes, New Jersey, USA) from the patients. Whole transcriptome sequencing (WTS) libraries were prepared using KAPA mRNA HyperPrep (Roche, Basel, Switzerland) or Stranded Total RNA Prep Ligation with Ribo-Zero Plus (Illumina, San Diego, California, USA) kits according to the manufacturer’s instructions.

The clinical significance of the variants was determined according to the criteria of the American College of Genomics and Genetics (ACMG) guidelines ^15,16^, and modifications ^17^. Variants in actionable genes, as defined by the ACMG, were searched in individuals who provided consent ^18^. Each variant classification was completed independently by two specialists.

We categorized the reports into positive, inconclusive, and negative. A positive result was defined by the presence of pathogenic/likely pathogenic variant(s) that fully or partially explained the patient’s phenotype. inconclusive reports featured variant(s) that did not define a molecular diagnosis, while negative reports indicated no genetic variant explaining the disorder.

We reported single nucleotide variants (SNVs), small indels (1-50 base pairs), copy number/structural variants (CNVs/SVs, more than 50 base pairs), and short tandem repeats (STRs). Both nuclear and mitochondrial DNA variants were investigated.

ExpansionHunter ^19^ was used to estimate STR copy numbers. Pre-mutation and full mutation expansion calls were confirmed by visual inspection of BAM reads. If necessary, orthogonal validation of STR calls was conducted.

We calculated the number of positive reports involving variants best captured by WGS, representing additional diagnostic gain over whole exome sequencing (WES). These included: CNVs/SVs involving fewer than three exons, deep intronic SNVs/small indels (25 nucleotides or more from the nearest exon), those in non-coding regions (regulatory regions, promoters, 5’ or 3’ untranslated regions – UTR), STR nucleotide expansions, and mitochondrial variants.

Global ancestry was estimated by principal component analysis (PCA) with aproximately 600,000 biallelic, autosomal variants from Illumina’s Infinium Global Screening Array (GSA) and the ADMIXTURE algorithm ^20^. Recent consanguinity estimates were performed by quantifying runs of homozygosity (NROH) with lengths ≥ 1 million base pairs (MB) and the sum of all NROH lengths (SROH) ^21^ (Supplementary Methods).

We are using the DROP tool ^22^ for WTS data, to guide the reassessment of Negative/Inconclusive reports based on candidate genes with aberrant expression or splicing (Supplementary Methods).

## 3. Results

### 3.1. Patients

We enrolled 8966 patients from 21 centers nationwide. The median age was 12 years (interquartile range, IQR = 5 - 32, minimum = 0, maximum = 92), with variations across disease cohorts (Supplementary Tables 1 and 2). Female participants comprised the majority of the overall sample (53.6%), with the highest proportion observed in the hereditary cancer risk syndromes cohort (87.9%) due to a history of breast cancer. Male participants represented 46.4% of the sample and predominated in the largest cohort (Neurologic disorders - 57.7%).

### 3.2. Diagnostic Yield

Among the enrolled patients, 7614 have already received whole genome sequencing (WGS) results. The overall diagnostic yield is 35.8% (2728/7614). Inconclusive reports amounted to 32.6% (2479/7614), while negative reports constituted 31.6% (2407/7614). The diagnostic yield varied according to disorder cohorts, ranging from 19.6% in the Immunologic disorders (229/1169) cohort to 74.3% in the Dermatologic disorders cohort (75/101). Among critically ill patients, the diagnostic yield was 34% (49/144). The Table 1 displays the number of enrolled patients and their respective diagnostic yields.

**Table 1:** Diagnostic yields.

| Cohort | Enrolled | Percent overall | Reports | Positive reports | Positive reports (with additional gains) | Diagnostic yield (%) | WGS yield gains (%) |
| --- | --- | --- | --- | --- | --- | --- | --- |
| Neurologic disorders | 3,027 | 33.76 | 2,459 | 940 | 79 | 38.2 | 8.4 |
| Neuromuscular disorders | 252 | 2.81 | 215 | 77 | 10 | 35.8 | 13.0 |
| Neurocutaneous disorders | 146 | 1.63 | 132 | 94 | 2 | 71.2 | 2.1 |
| Inborn errors of metabolism | 392 | 4.37 | 342 | 157 | 13 | 45.9 | 8.3 |
| Skeletal disorders | 350 | 3.90 | 302 | 177 | 5 | 58.6 | 2.8 |
| Hereditary cancer risk syndromes | 1,553 | 17.32 | 1,352 | 340 | 11 | 25.1 | 3.2 |
| Clinically recognizable genetic syndromes | 537 | 5.99 | 469 | 227 | 8 | 48.4 | 3.5 |
| Endocrine disorders | 307 | 3.42 | 242 | 85 | 4 | 35.1 | 4.7 |
| Connective tissue disorders | 142 | 1.58 | 124 | 54 | 1 | 43.5 | 1.9 |
| Immunologic disorders | 1,328 | 14.81 | 1,169 | 229 | 7 | 19.6 | 3.1 |
| Ophthalmologic disorders | 154 | 1.72 | 123 | 57 | 2 | 46.3 | 3.5 |
| Hearing or ear disorders | 42 | 0.47 | 36 | 17 | 2 | 47.2 | 11.8 |
| Renal and urinary tract disorders | 106 | 1.18 | 87 | 44 | - | 50.6 | - |
| Dermatologic disorders | 119 | 1.33 | 101 | 75 | 2 | 74.3 | 2.7 |
| Hematologic disorders | 116 | 1.29 | 91 | 37 | 3 | 40.7 | 8.1 |
| Cardiological disorders | 93 | 1.04 | 86 | 25 | - | 29.1 | - |
| Pulmonary disorders | 58 | 0.65 | 52 | 23 | 1 | 44.2 | 4.3 |
| Gastroenterologic disorders | 97 | 1.08 | 88 | 21 | 1 | 23.9 | 4.8 |
| Vascular disorders | 3 | 0.03 | - | - | - | - | - |
| Critically ill patients | 144 | 1.61 | 144 | 49 | 3 | 34.0 | 6.1 |
| Overall | 8,966 | 100.00 | 7,614 | 2,728 | 154 | 35.8 | 5.6 |

### 3.3. Ancestry and consanguinity

We assessed the individual ancestry background by PCA and the ADMIXTURE algorithm ^20^ with 600,000 biallelic variants. We observed there is a remarkable variation regarding ancestry backgrounds in our sample, as expected. Individuals from the GRAR sample had significant proportion of EUR background (median = 74.7%, IQR = 62.5% - 86.3%), followed by AFR contribution (median = 23.8%, IQR = 12.3% - 36.4%).

The PCA results also confirm the strong admixture. The Brazilian samples do not tightly cluster around the reference samples, instead behaving like a gradient (Figure 1).

**Figure 1.**
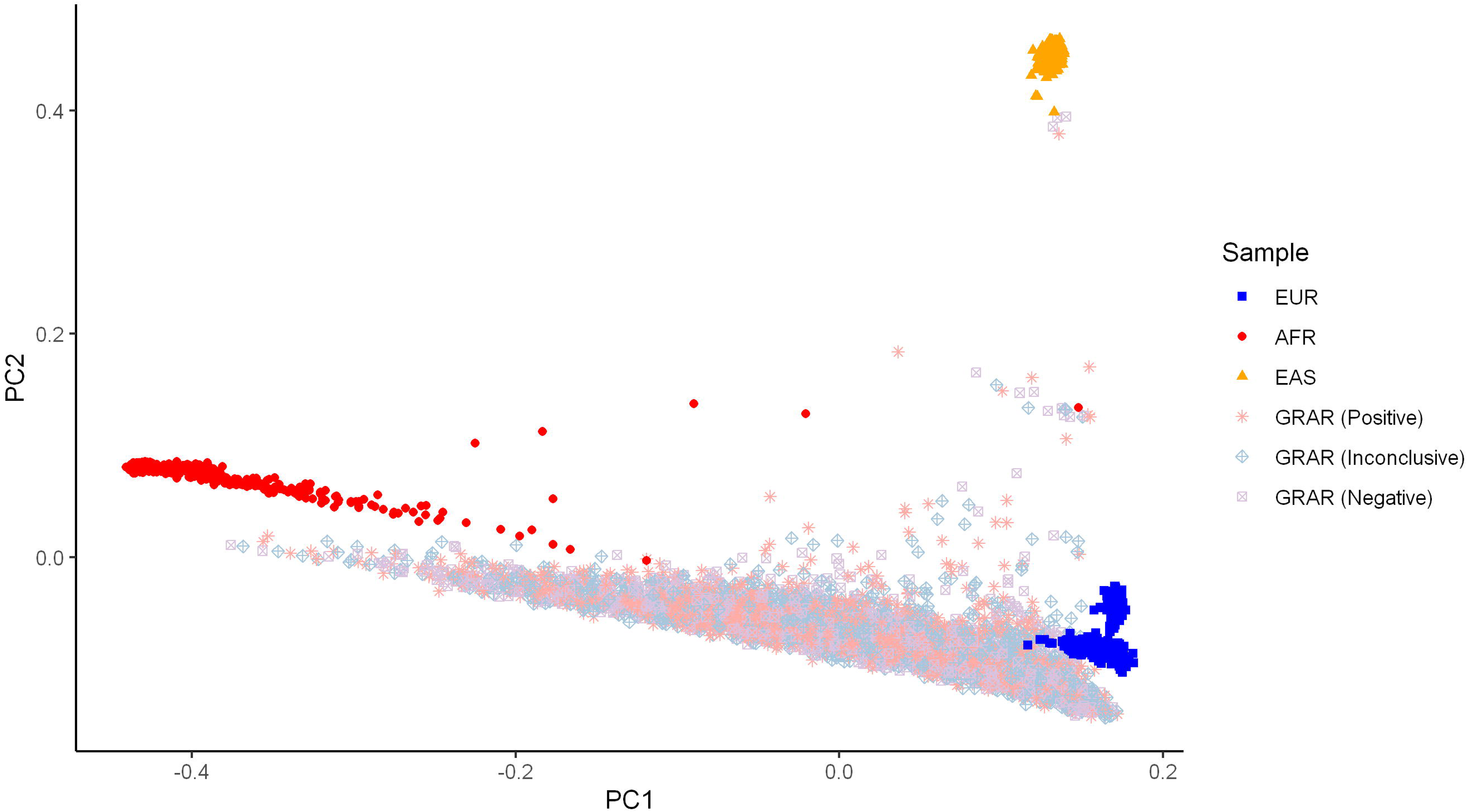
Principal Component Analysis (PCA) plot with reference populations from the 1000 Genomes Project representing the ancestry backgrounds in the individuals recruited by The Brazilian Rare Genomes Project (BRGP). Colors and point shapes represent samples and, in the case of BRGP samples, the report result. PC: principal component. The PCA was performed with approximately 600,000 biallelic variants.

**Figure 2.**
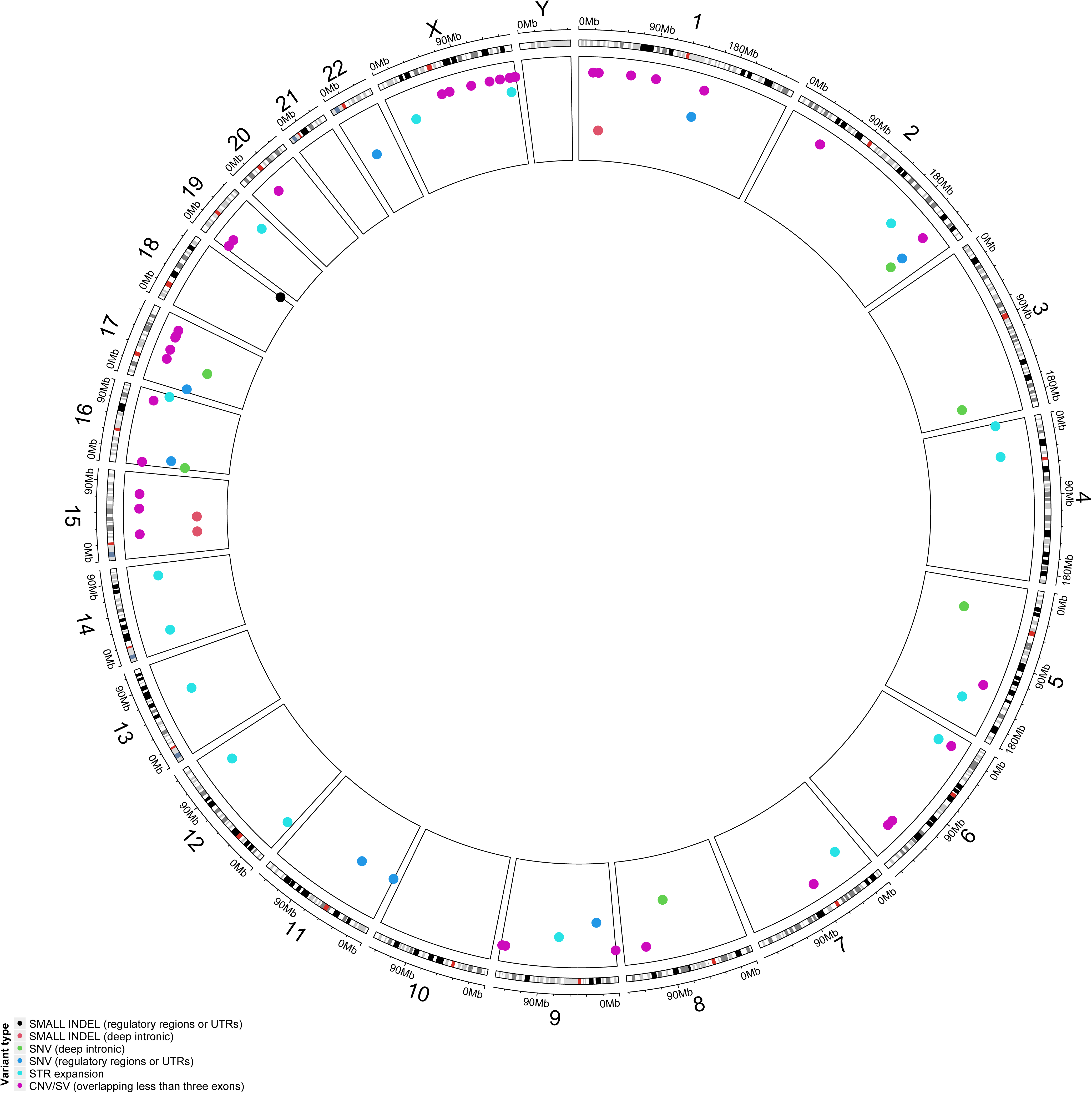
Circular plot representing the nuclear genome. Points represent location of pathogenic/likely pathogenic variants that are best captured by WGS, thus represent additional diagnostic gain over WES.

The median SROH in the GRAR sample was relatively low (6.2 MB, IQR = 3.3 MB - 11.8 MB), meaning that only a relevant minority of patients presented strong evidence for recent consanguinity, with few patients presenting extreme SROH values. In fact, according to Matalonga et al.’s ^21^ SROH thresholds, just 165 (2.2%) individuals were probably consanguineous and 558 (7.3%) were consanguineous. Thus, at most 9.5% of the individuals from our sample present evidence for consanguinity.

### 3.4. Affected genes

A total of 2488 genes with records in the Online Mendelian Inheritance in Man (OMIM) database ^23^ presented variants among the 2728 patients with positive reports. These genes were associated with diseases exhibiting the following modes of inheritance: autosomal recessive (AR): 47.1%; autosomal dominant (AD): 29.7%; X-linked recessive (XLR): 5.3%; AR or AD: 5.3%; X-linked dominant (XLD): 1.2%; mitochondrial: 0.8%; XLR or XLD: 0.4%; X-linked inheritance: 0.6%; and a minority of genes with no inheritance data available: 9.5%.

The Supplementary Table 3 presents a list of genes that appeared in positive reports, ranked by their frequency of occurrence.

We submitted selected variant interpretations (excluding secondary findings) to the ClinVar database. The submissions included a total of 1891 interpretations: 474 (25.1%) pathogenic, 473 (25.0%) likely pathogenic and 944 (49.9%) variants of uncertain significance (VUS). These interpretations were distributed across 971 unique genes.

### 3.5. Additional diagnostic gains through WGS

Among the 2728 individuals with positive WGS reports, WGS yielded additional diagnostic gains for 154 (5.6%) of them, by providing information regarding variants less likely to be detected by other methodologies. This resulted in 5.6% more conclusive reports compared to WES.

The advantage of WGS varied according to disorder cohorts, ranging between 1.9% in the Connective tissue disorders cohort and 13% in the Neuromuscular disorders cohort. The Critically ill cohort exhibited a 6.1% increase. The Table 1 also summarizes the additional gains per cohort.

Short CNV/SV involving fewer than three exons had a significant prevalence: they were identified in 48 among the 471 positive patients with pathogenic/likely pathogenic CNV/SV events (10.2%). Notably, 20.8% (10/48) of the short structural variants were located on the X chromosome. Consequently, WGS in our setting contributed to a slight increase in the diagnostic yield of X-linked conditions, such as neurodevelopmental disorders (e.g., *AFF2*, *ZC4H2*), immunodeficiencies (e.g., *BTK*, *XIAP*), and inborn errors of metabolism (e.g., *IDS*).

Our WGS protocol provided positive reports involving STR expansions for 41 patients. The majority of STR expansion variants were associated with neurological conditions (38 cases, 92.7%), except for *HOXD13*: two cases (4.9%); *HOXA13*: one case (2.4%). Genes with the most detected STR expansion variants detected included: *FMR1*: 10 cases; *ATXN8OS*: seven cases; *ATXN1*, *ATXN3*, and *DMPK*: three cases each; *ARX*, *FXN*, and *PABPN1*: two cases each; and *ATN1*, *ATXN2*, *HTT*, *JPH3*, *PHOX2B*, and *PPP2R2B*: one case each. STR expansion cases accounted for 1.5% of all positive cases.

Additionally, 38 probands received a positive mitochondrial disorder diagnosis due to variants in the mitochondrial genome (23 with SNVs, 7 with small indels, and 8 with rare mitochondrial CNV/SV). WGS provided very high mitochondrial read coverage, with median coverage 3874.1X (IQR = 3048.1X - 4784.3X, minimum = 937.7X, maximum = 12934.5X), which facilitated the identification of those structural variants.

The remaining 25 patients had miscellaneous SNVs and small indels in non-coding regions (deep intronic, regulatory or UTR regions). The Table 2 displays the reported events, stratified by variant type. The Figure 1 illustrates the location of nuclear variants categorized as WGS additional gains.

**Table 2:**
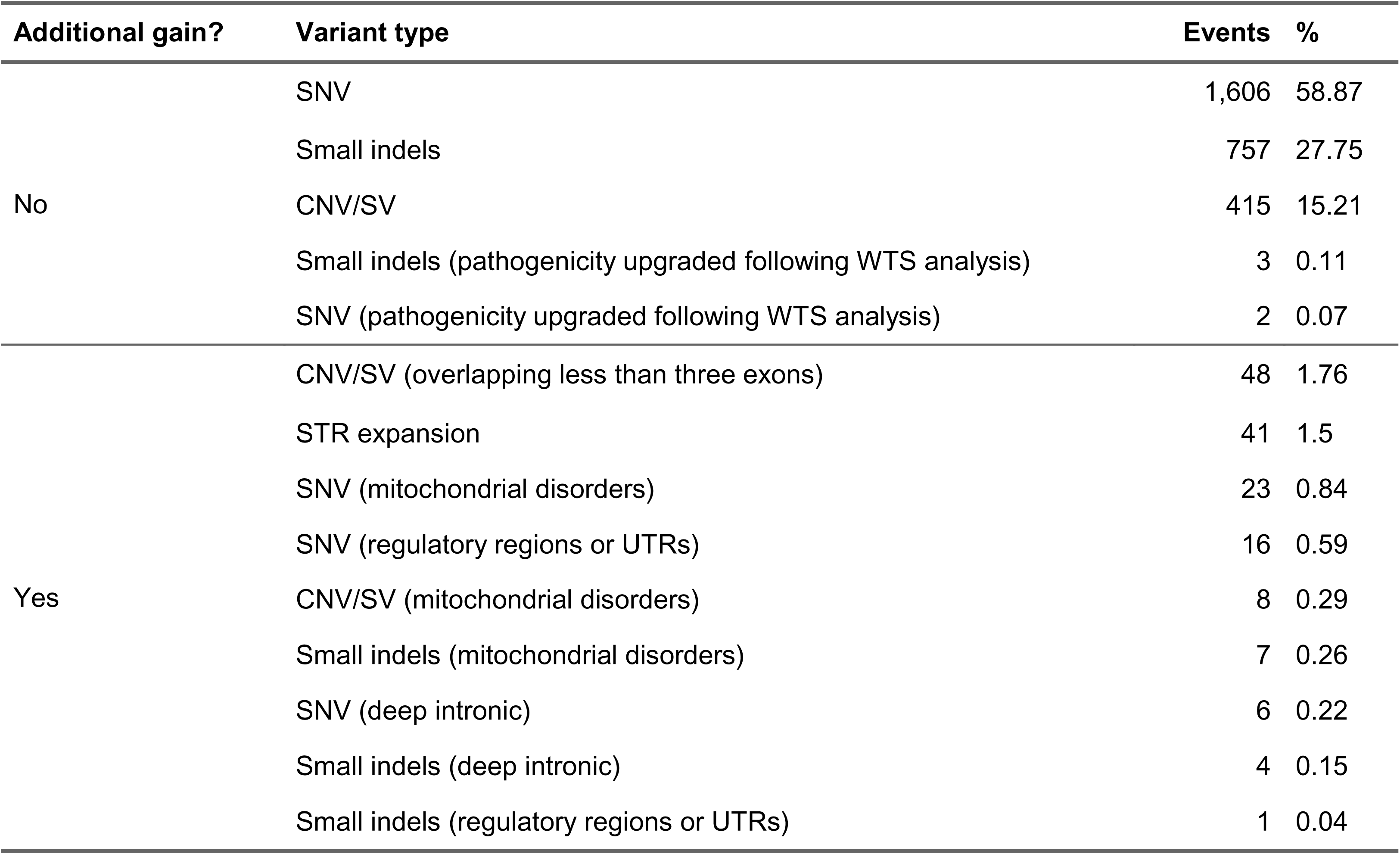
Events in positive reports, stratified according to additional gain from whole genome sequencing and variant type.

### 3.6. Transcriptomics: contribution to diagnosis

At present, we analyzed 1025 WTS. Currently, 48 of them have been analyzed. The DROP tool results aided in the reinterpretation of five reports following the upgrade of their ACMG classification criteria in the light of the new evidence brought by the detection of aberrant expression or the visualization of aberrant splicing events. They represent 10.4% among the analyzed transcriptomes and 0.2% of all positive cases. The Supplementary Table 4 presents a summary of the cases solved with the help of RNA-Seq analysis.

## 4. Discussion

The Brazilian Rare Genomes Project is currently the largest initiative Brazil focused on diagnosing rare diseases and hereditary cancer risk syndromes. With over 7600 reports released, the project assists individuals and their families across the country in understanding their genetic conditions.

Most patients involved have not undergone prior genetic testing, unlike the participants in Genomic England’s 100,000 Genomes Project ^24^, where some patients have previous WES or other tests before WGS. The BRGP has observed a diagnostic yield of 35.8%, which aligns with estimates from other studies. For instance, Genomic England reported a yield of 33% among patients without previous testing.

A meta-analysis of 37 studies involving more than 20,000 children who underwent WGS suggested an overall diagnostic yield of 41% (95% CI = 34% - 48%) ^25^, while other studies found yields as high as 68.3% ^26^, depending on the population and disorder cohort.

The 100,000 Genomes project indicated that 14% of diagnostics represented additional gains by WGS compared to other diagnostic tests, involving variants in noncoding, structural, and mitochondrial genome regions. Similar benefits are noted in our study. There is demonstrated value in using WGS as the initial diagnostic test for rare diseases patients, particularly for heterogeneous and hard-to-diagnose disorder cohorts such as immunologic disorders. A study of primary immunodeficiency patients identified 69 novel variants, including eight structural variants undetectable by WES ^27^. The Broad Center for Mendelian Genomics reported that 8.2% of their findings involved variants ascertainable only through WGS ^28^.

Our estimates correspond with previous studies, showing diagnostic yields between 5.6% and 10.2% among various patient groups.

We have implemented a WGS workflow to expedite reporting for critically ill neonates in neonatal/pediatric intensive care units (NICU/PICU), with a diagnostic yield of 34.0%, reflecting international settings that range between 21% and 46% ^29–32^.

Our efforts contribute to the scientific community: with over 1900 interpretations deposited in ClinVar, providing new data for clinical interpretations globally. This aligns with current movements to increase representation from diverse populations in genomic databases ^33,34^.

To reassess inconclusive or negative reports, we have whole transcriptome sequencing data from over 1,000 patients and long-read sequencing data from over 200 patients. Previous studies indicate RNA-Seq-based diagnostic yields between 3% ^35^ and 16% ^36^, consistent with our current proportion of 10.4%. Long-read sequencing is reassessing poorly covered genome regions, enabling epigenomics analyses as well.

In conclusion, the BRGP showcases that WGS and WTS are essential tools for diagnosing rare diseases and hereditary cancer risk syndromes. WGS improves diagnostic yield by capturing variants missed by traditional tests, with rates ranging from 35.8% to 10.4% among different patient groups. WTS identifies aberrant expression and splicing events, aiding in the reinterpretation of existing reports. This method has resolved 10.4% of our analyzed transcriptomes of patients with specific genomic findings. Together, WGS and WTS enhance diagnostic accuracy, uncover genetic conditions, and support research and clinical practice.

## Data Availability

All data produced in the present work are contained in the manuscript.

