## Supplementary Methods for "Whole Genome Sequencing for the Diagnosis of Rare Disorders"

### 1.1. Global ancestry and admixture

We aggregated GVCF files into a multisample VCF file (msVCF) also through DRAGEN. We developed a WDL/Cromwell pipeline (named Rocket-joint, <https://github.com/GenesisGenomics/rocket-joint>) to merge the GVCF files through processing in the AWS cloud. We acknowledge Illumina’s DRAGEN Staff for kindly providing the code for GVCF merging that served as the basis for the development of Rocket-joint.

Finally, the msVCF was imported into Hail ^1^ in AWS Elastic MapReduce (EMR) clusters. Through Hail, we performed sample- and variant-level quality control, by removing both samples and variants with call rate < 85%, variants with transition/transversion (Ti/Tv) ratio < 1.9, and any variants not passing DRAGEN’s default hard-filters. Next, we merged our dataset with a 1000 Genomes Project ^2^ reference population dataset to run a Principal Components Analysis (PCA) for the estimation of the first 10 ancestry-related principal components based on aproximately 600,000 biallelic, autosomal variants contained in the Illumina’s Infinium Global Screening Array (GSA).

Besides the PCA, we estimated global ancestry proportions through ADMIXTURE algorithm ^3^ with the same GSA alleles and K = 3, since the Brazilian population is mostly three-way admixed ^4^. We used the EUR and AFR superpopulations from the 1000 Genomes Project ^2^ to serve as references for the European and African components, respectively. Since we unfortunately did not have Amerindian datasets available, we used the East Asian (EAS) superpopulation as a proxy to account for the third component.

### 1.2. Consanguinity

In parallel, we used DRAGEN’s outputs to quantify the number of runs of homozygosity (NROH) with lengths ≥ 1 million base pairs (MB), the sum of all NROH lengths (SROH), and the frequency of ROH (FROH) across the genome of each sample through scripts for R software version 4.4.1. We used Matalonga et al.’s ^5^ SROH thresholds to summarize evidence for recent consanguinity: no consanguinity: SROH ≤ 22 MB; probable non-consanguinity: SROH > 22 MB and SROH < 79 MB; probable consanguinity: SROH ≥ 79 MB and SROH ≤ 123 MB; consanguinity: SROH > 123 MB.

### 1.3. RNA-Seq

We extracted whole RNA from peripheral blood mononuclear cells (PBMCs). We prepared whole transcriptome sequencing (WTS) libraries with KAPA mRNA HyperPrep (Roche, Basel, Switzerland) or Stranded Total RNA Prep Ligation with Ribo-Zero Plus (Illumina, San Diego, California, United States) kits according to the manufacturer’s instructions. The RNA-Seq was performed in Illumina’s NextSeq 6000 platform. The sequencing reads were mapped to the reference genome (GRCh38 build) with DRAGEN version 3.8.4 with the following parameter values: COUNT_MODE = “IntersectionStrict”, STRAND = “no”, COUNT_OVERLAPS = True, PAIRED_END = True.

The BAM files were processed by the DROP tool version 1.1.1 ^6^. We executed all three DROP modules: OUTRIDER (aberrant expression) ^7^, FRASER (aberrant splicing) ^8^, and MAE (monoallelic expression). The last one required VCF files obtained via the WGS from the same patients, which were imported into DROP accordingly.

We are currently using the DROP tool ^6^ to guide the reassessment of Negative/Inconclusive reports based on candidate genes with aberrant expression or splicing.

##### References

1.	Hail Team. Hail version 0.2. (<https://github.com/hail-is/hail>).
