## Supplementary Table 1 for "Whole Genome Sequencing for the Diagnosis of Rare Disorders"

Supplementary Table 1: Enrolled patients' characteristics (stratified by disorders cohorts).

| Cohort | Sex | All patients, n (%) | Neonates and infants, n (%) | Other pediatric patients, n (%) | Adult patients, n (%) |
| --- | --- | --- | --- | --- | --- |
| Neurologic disorders | Female | 1281 (42.3) | 175 (52.2) | 881 (39.9) | 225 (46.3) |
|  | Male | 1746 (57.7) | 160 (47.8) | 1325 (60.1) | 261 (53.7) |
|  | Total | 3027 (100) | 335 (100) | 2206 (100) | 486 (100) |
| Neuromuscular disorders | Female | 126 (50) | 15 (50) | 62 (48.4) | 49 (52.1) |
|  | Male | 126 (50) | 15 (50) | 66 (51.6) | 45 (47.9) |
|  | Total | 252 (100) | 30 (100) | 128 (100) | 94 (100) |
| Neurocutaneous disorders | Female | 73 (50) | 3 (42.9) | 31 (38.3) | 39 (67.2) |
|  | Male | 73 (50) | 4 (57.1) | 50 (61.7) | 19 (32.8) |
|  | Total | 146 (100) | 7 (100) | 81 (100) | 58 (100) |
| Inborn errors of metabolism | Female | 184 (46.9) | 40 (41.7) | 109 (47) | 35 (54.7) |
|  | Male | 208 (53.1) | 56 (58.3) | 123 (53) | 29 (45.3) |
|  | Total | 392 (100) | 96 (100) | 232 (100) | 64 (100) |
| Skeletal disorders | Female | 173 (49.4) | 18 (34) | 120 (51.5) | 35 (54.7) |
|  | Male | 177 (50.6) | 35 (66) | 113 (48.5) | 29 (45.3) |
|  | Total | 350 (100) | 53 (100) | 233 (100) | 64 (100) |
| Hereditary cancer risk syndromes | Female | 1365 (87.9) | 4 (57.1) | 17 (48.6) | 1344 (88.9) |
|  | Male | 188 (12.1) | 3 (42.9) | 18 (51.4) | 167 (11.1) |
|  | Total | 1553 (100) | 7 (100) | 35 (100) | 1511 (100) |
| Clinically recognizable genetic syndromes | Female | 262 (48.8) | 44 (44.9) | 172 (48.2) | 46 (56.1) |
|  | Male | 275 (51.2) | 54 (55.1) | 185 (51.8) | 36 (43.9) |
|  | Total | 537 (100) | 98 (100) | 357 (100) | 82 (100) |
| Endocrine disorders | Female | 136 (44.3) | 20 (38.5) | 83 (43.9) | 33 (50) |
|  | Male | 171 (55.7) | 32 (61.5) | 106 (56.1) | 33 (50) |
|  | Total | 307 (100) | 52 (100) | 189 (100) | 66 (100) |
| Connective tissue disorders | Female | 74 (52.1) | 2 (100) | 44 (47.3) | 28 (59.6) |
|  | Male | 68 (47.9) | 0 (0) | 49 (52.7) | 19 (40.4) |
|  | Total | 142 (100) | 2 (100) | 93 (100) | 47 (100) |
| Immunologic disorders | Female | 673 (50.7) | 39 (39.4) | 303 (45.4) | 331 (58.9) |
|  | Male | 655 (49.3) | 60 (60.6) | 364 (54.6) | 231 (41.1) |
|  | Total | 1328 (100) | 99 (100) | 667 (100) | 562 (100) |
| Ophthalmologic disorders | Female | 74 (48.1) | 17 (48.6) | 32 (42.7) | 25 (56.8) |
|  | Male | 80 (51.9) | 18 (51.4) | 43 (57.3) | 19 (43.2) |
|  | Total | 154 (100) | 35 (100) | 75 (100) | 44 (100) |
| Hearing or ear disorders | Female | 20 (47.6) | 1 (33.3) | 14 (53.8) | 5 (38.5) |
|  | Male | 22 (52.4) | 2 (66.7) | 12 (46.2) | 8 (61.5) |
|  | Total | 42 (100) | 3 (100) | 26 (100) | 13 (100) |
| Renal and urinary tract disorders | Female | 52 (49.1) | 5 (35.7) | 37 (47.4) | 10 (71.4) |
|  | Male | 54 (50.9) | 9 (64.3) | 41 (52.6) | 4 (28.6) |
|  | Total | 106 (100) | 14 (100) | 78 (100) | 14 (100) |
| Dermatologic disorders | Female | 64 (53.8) | 12 (48) | 30 (47.6) | 22 (71) |
|  | Male | 55 (46.2) | 13 (52) | 33 (52.4) | 9 (29) |
|  | Total | 119 (100) | 25 (100) | 63 (100) | 31 (100) |
| Hematologic disorders | Female | 67 (57.8) | 9 (64.3) | 33 (50.8) | 25 (67.6) |
|  | Male | 49 (42.2) | 5 (35.7) | 32 (49.2) | 12 (32.4) |
|  | Total | 116 (100) | 14 (100) | 65 (100) | 37 (100) |
| Cardiological disorders | Female | 42 (45.2) | 12 (48) | 20 (44.4) | 10 (43.5) |
|  | Male | 51 (54.8) | 13 (52) | 25 (55.6) | 13 (56.5) |
|  | Total | 93 (100) | 25 (100) | 45 (100) | 23 (100) |
| Pulmonary disorders | Female | 35 (60.3) | 3 (60) | 28 (59.6) | 4 (66.7) |
|  | Male | 23 (39.7) | 2 (40) | 19 (40.4) | 2 (33.3) |
|  | Total | 58 (100) | 5 (100) | 47 (100) | 6 (100) |
| Gastroenterologic disorders | Female | 39 (40.2) | 6 (31.6) | 30 (42.3) | 3 (42.9) |
|  | Male | 58 (59.8) | 13 (68.4) | 41 (57.7) | 4 (57.1) |
|  | Total | 97 (100) | 19 (100) | 71 (100) | 7 (100) |
| Vascular disorders | Female | 1 (33.3) | 0 (100) | 1 (33.3) | 0 (100) |
|  | Male | 2 (66.7) | 0 (100) | 2 (66.7) | 0 (100) |
|  | Total | 3 (100) | 0 (100) | 3 (100) | 0 (100) |
| Critically ill patients | Female | 64 (44.4) | 64 (44.4) | 0 (100) | 0 (100) |
|  | Male | 80 (55.6) | 80 (55.6) | 0 (100) | 0 (100) |
|  | Total | 144 (100) | 144 (100) | 0 (100) | 0 (100) |
