## Supplementary Table 2 for "Whole Genome Sequencing for the Diagnosis of Rare Disorders"

Supplementary Table 2: Enrolled patients' age group distribution (stratified by disorders cohorts).

|  | **Age group in years, n (%)** | | | | | | | | | | | | | | | | |
| --- | --- | --- | --- | --- | --- | --- | --- | --- | --- | --- | --- | --- | --- | --- | --- | --- | --- |
| **Cohort** | **0-4** | **5-9** | **10-14** | **15-19** | **20-24** | **25-29** | **30-34** | **35-39** | **40-44** | **45-49** | **50-54** | **55-59** | **60-64** | **65-69** | **70-74** | **75-79** | **80+** |
| Neurologic disorders | 884 (29.2) | 782 (25.83) | 563 (18.6) | 353 (11.66) | 139 (4.59) | 67 (2.21) | 46 (1.52) | 39 (1.29) | 33 (1.09) | 23 (0.76) | 28 (0.93) | 29 (0.96) | 15 (0.5) | 15 (0.5) | 7 (0.23) | 4 (0.13) | 0 (0) |
| Neuromuscular disorders | 61 (24.21) | 47 (18.65) | 27 (10.71) | 27 (10.71) | 3 (1.19) | 10 (3.97) | 10 (3.97) | 14 (5.56) | 6 (2.38) | 10 (3.97) | 8 (3.17) | 10 (3.97) | 14 (5.56) | 3 (1.19) | 1 (0.4) | 1 (0.4) | 0 (0) |
| Neurocutaneous disorders | 27 (18.49) | 24 (16.44) | 22 (15.07) | 16 (10.96) | 14 (9.59) | 11 (7.53) | 6 (4.11) | 9 (6.16) | 3 (2.05) | 3 (2.05) | 5 (3.42) | 3 (2.05) | 0 (0) | 1 (0.68) | 2 (1.37) | 0 (0) | 0 (0) |
| Inborn errors of metabolism | 169 (43.11) | 84 (21.43) | 46 (11.73) | 41 (10.46) | 17 (4.34) | 8 (2.04) | 4 (1.02) | 7 (1.79) | 7 (1.79) | 1 (0.26) | 3 (0.77) | 2 (0.51) | 1 (0.26) | 1 (0.26) | 1 (0.26) | 0 (0) | 0 (0) |
| Skeletal disorders | 102 (29.14) | 80 (22.86) | 76 (21.71) | 30 (8.57) | 18 (5.14) | 14 (4) | 7 (2) | 3 (0.86) | 5 (1.43) | 6 (1.71) | 8 (2.29) | 0 (0) | 0 (0) | 0 (0) | 1 (0.29) | 0 (0) | 0 (0) |
| Hereditary cancer risk syndromes | 14 (0.9) | 10 (0.64) | 5 (0.32) | 17 (1.09) | 33 (2.12) | 87 (5.6) | 168 (10.82) | 242 (15.58) | 274 (17.64) | 217 (13.97) | 138 (8.89) | 133 (8.56) | 84 (5.41) | 70 (4.51) | 31 (2) | 24 (1.55) | 6 (0.39) |
| Clinically recognizable genetic syndromes | 190 (35.38) | 117 (21.79) | 92 (17.13) | 61 (11.36) | 37 (6.89) | 16 (2.98) | 7 (1.3) | 8 (1.49) | 3 (0.56) | 3 (0.56) | 1 (0.19) | 1 (0.19) | 0 (0) | 0 (0) | 1 (0.19) | 0 (0) | 0 (0) |
| Endocrine disorders | 92 (29.97) | 65 (21.17) | 52 (16.94) | 36 (11.73) | 17 (5.54) | 10 (3.26) | 15 (4.89) | 8 (2.61) | 2 (0.65) | 3 (0.98) | 3 (0.98) | 2 (0.65) | 2 (0.65) | 0 (0) | 0 (0) | 0 (0) | 0 (0) |
| Connective tissue disorders | 16 (11.27) | 20 (14.08) | 32 (22.54) | 29 (20.42) | 12 (8.45) | 7 (4.93) | 4 (2.82) | 7 (4.93) | 6 (4.23) | 7 (4.93) | 2 (1.41) | 0 (0) | 0 (0) | 0 (0) | 0 (0) | 0 (0) | 0 (0) |
| Immunologic disorders | 273 (20.56) | 200 (15.06) | 199 (14.98) | 107 (8.06) | 87 (6.55) | 67 (5.05) | 75 (5.65) | 81 (6.1) | 56 (4.22) | 46 (3.46) | 39 (2.94) | 37 (2.79) | 30 (2.26) | 16 (1.2) | 8 (0.6) | 3 (0.23) | 4 (0.3) |
| Ophthalmologic disorders | 59 (38.31) | 18 (11.69) | 22 (14.29) | 11 (7.14) | 7 (4.55) | 10 (6.49) | 3 (1.95) | 7 (4.55) | 7 (4.55) | 3 (1.95) | 2 (1.3) | 3 (1.95) | 1 (0.65) | 1 (0.65) | 0 (0) | 0 (0) | 0 (0) |
| Hearing or ear disorders | 10 (23.81) | 7 (16.67) | 8 (19.05) | 5 (11.9) | 1 (2.38) | 3 (7.14) | 1 (2.38) | 2 (4.76) | 0 (0) | 2 (4.76) | 0 (0) | 2 (4.76) | 1 (2.38) | 0 (0) | 0 (0) | 0 (0) | 0 (0) |
| Renal and urinary tract disorders | 30 (28.3) | 20 (18.87) | 24 (22.64) | 18 (16.98) | 4 (3.77) | 2 (1.89) | 2 (1.89) | 2 (1.89) | 1 (0.94) | 1 (0.94) | 1 (0.94) | 1 (0.94) | 0 (0) | 0 (0) | 0 (0) | 0 (0) | 0 (0) |
| Dermatologic disorders | 42 (35.29) | 30 (25.21) | 7 (5.88) | 12 (10.08) | 9 (7.56) | 8 (6.72) | 2 (1.68) | 2 (1.68) | 3 (2.52) | 2 (1.68) | 1 (0.84) | 0 (0) | 0 (0) | 0 (0) | 1 (0.84) | 0 (0) | 0 (0) |
| Hematologic disorders | 25 (21.55) | 18 (15.52) | 28 (24.14) | 9 (7.76) | 6 (5.17) | 6 (5.17) | 3 (2.59) | 3 (2.59) | 6 (5.17) | 4 (3.45) | 2 (1.72) | 2 (1.72) | 1 (0.86) | 1 (0.86) | 2 (1.72) | 0 (0) | 0 (0) |
| Cardiological disorders | 40 (43.01) | 11 (11.83) | 13 (13.98) | 7 (7.53) | 3 (3.23) | 5 (5.38) | 1 (1.08) | 4 (4.3) | 2 (2.15) | 1 (1.08) | 1 (1.08) | 1 (1.08) | 1 (1.08) | 0 (0) | 2 (2.15) | 0 (0) | 1 (1.08) |
| Pulmonary disorders | 14 (24.14) | 13 (22.41) | 17 (29.31) | 10 (17.24) | 2 (3.45) | 0 (0) | 1 (1.72) | 1 (1.72) | 0 (0) | 0 (0) | 0 (0) | 0 (0) | 0 (0) | 0 (0) | 0 (0) | 0 (0) | 0 (0) |
| Gastroenterologic disorders | 41 (42.27) | 22 (22.68) | 16 (16.49) | 11 (11.34) | 1 (1.03) | 1 (1.03) | 0 (0) | 1 (1.03) | 2 (2.06) | 0 (0) | 1 (1.03) | 1 (1.03) | 0 (0) | 0 (0) | 0 (0) | 0 (0) | 0 (0) |
| Vascular disorders | 1 (33.33) | 2 (66.67) | 0 (0) | 0 (0) | 0 (0) | 0 (0) | 0 (0) | 0 (0) | 0 (0) | 0 (0) | 0 (0) | 0 (0) | 0 (0) | 0 (0) | 0 (0) | 0 (0) | 0 (0) |
| Critically ill patients | 144 (100) | 0 (0) | 0 (0) | 0 (0) | 0 (0) | 0 (0) | 0 (0) | 0 (0) | 0 (0) | 0 (0) | 0 (0) | 0 (0) | 0 (0) | 0 (0) | 0 (0) | 0 (0) | 0 (0) |
