## Supplementary Table 4 for "Whole Genome Sequencing for the Diagnosis of Rare Disorders"

Supplementary Table 4: Summary of the cases solved with the help of RNA-Seq analysis.

| Variant type | Gene | RefSeq ID | HGVS c. | HGVS p. | Region | Case description |
| --- | --- | --- | --- | --- | --- | --- |
| Small indel | *SLC10A7* | NM_001029998.6 | c.471+3_471+6del |  | - | A 6-year-old female patient was under investigation for a clinical phenotype characterized by disproportionate short stature, shortening of long bones, ligamentous laxity, femoral epiphysis hypoplasia, genu valgum, and pes planus. Whole genome sequencing (WGS) identified two homozygous variants of uncertain significance, potentially associated with the patient’s phenotype: *SLC10A7* (NM_001029998.6):c.471+3_471+6del and *CFAP410* (NM_004928.3):c.78-547T>C. Both variants are extremely rare in the gnomAD 4.0 population database and were predicted in silico to affect exon splicing using the SpliceAI tool (ref). To assess their functional impact, transcriptome sequencing was performed. The analysis revealed that the *CFAP410*:c .78-547T>C variant had no detectable effect on RNA splicing in peripheral blood. In contrast, the SLC10A7:c .471+3_471+6del variant resulted in aberrant splicing, leading to the complete skipping of exon 6. Although this alteration does not disrupt the protein’s reading frame, it occurs in a critical functional region of the protein. Based on these findings and in accordance with the latest recommendations of the ClinGen SVI Splicing Subgroup ^1^, the *SLC10A7* variant was reclassified as likely pathogenic. These findings demonstrate the clinical utility of transcriptome analysis in elucidating the functional impact of genetic variants, enabling accurate diagnosis and informed management for patients with complex clinical presentations. |
| Uniparental disomy | *SNRNP* | NM_021177.5 |  |  | 15q11-15q13 | A 9-year-old boy presenting obesity, hyperphagia, and neurodevelopmental delay had his whole genome analyzed. At first, two VUS were reported on *PHIP* and *MKKS* genes only, leading to an Inconclusive report. After the boy's transcriptome was sequenced, we noticed that *SNRNP* expression was remarkedly reduced compared to other samples sequenced in the same batch. This observation led to additional analyses that showed reduced heterozygosity on chromosome 15q11-15q13 region, which overlaps the *SNRNP* locus, suggesting a segmental uniparental disomy event. Additionally, we resequenced the patient sample with long-read sequencing (Oxford Nanopore Technologies library). The methylation analysis revealed hypermethylation in the *SNRNP* locus, reinforcing the disomy hypothesis. Thus, all this evidence led to a diagnosis of Prader-Willi syndrome. |
| Small indel | *MSH3* | NM_002439.5 | c.2318+5del |  | - | A case of familial adenomatous polyposis yielded an inconclusive result, with a homozygous variant identified in the *MSH3* gene: a one-base-pair deletion at the splicing site (NM_002439.5:c.2318+5del). Initially reported as a variant of uncertain significance, the in-silico splice site analysis predicted that proper splicing was affected, and the variant has an extremely low frequency in general population database. This variant has also been submitted to ClinVar with conflicting classifications, though it has never been documented in the literature. To further investigate its impact, we performed transcriptome sequencing, which confirmed that this deletion leads to abnormal splicing, resulting in exon 16 skipping. This alteration causes an out-of-frame consequence predicted to trigger nonsense-mediated decay. These findings are supported by other reports described in ClinVar submissions. Based on these findings and following the latest classification guidelines ^1^, this variant was reclassified as likely pathogenic. |
| Small indel | *STAT5B* | NM_012448.4 | c.424_427del  c.90G>A | p.(Leu142Argfs*20)  p.(Val30=) | - | A 22-year-old woman, presenting recurrent infections, interstitial pneumonitis, arthritis, and growth delay had her genome analyzed. Our experts detected two heterozygous variants in the *STAT5B* gene: one was a small indel pathogenic variant that led to frameshift (NM_012448.4:c.424_427del, NP_036580.2:p.(L142Rfs*20)), and the other was a synonymous SNV (NM_012448.4:c.90G>A, NP_036580.2:p.(Val30=)), that was classified as VUS. The phase of the variants could not be defined. After her transcriptome was sequenced, we observed that *STAT5B* expression was reduced compared to other samples, and we visualized an aberrant splicing event that potentially rendered a frameshift as well. Therefore, we suppose that the two variants are actually in trans. Possibly one or both of the frameshifts trigger the nonsense-mediated mRNA decay pathway, which would explain the reduced expression. Thus, we upgraded the synonymous variant classification to pathogenic, leading to a possible diagnosis of growth hormone insensitivity syndrome with immune dysregulation. |
| SNV | *DDX3X* | NM_001193416.3 | c.1769G>A | p.(Ser590Asn) | - | A 6-year-old girl presenting speech acquisition delay, dysmorphic features, cognitive impairment, and hyperactivity had her genome analyzed. Our experts reported three heterozygous missense SNVs in the genes: *DDX3X*, *BRPF1*, and *KAT6A*, and all three were initially classified as VUS. The girl transcriptome revealed an aberrant splicing event in the *DDX3X* transcript: an elongation of the exon 15. Thus, the experts reclassified the *DDX3X* variant (NM_001193416.3:c.1769G>A, NP_001180345.1:p.(Ser590Asn)) as likely pathogenic, leading to a tentative diagnosis of X-linked intellectual developmental disorder, Snijders Blok type. |

1.	Walker LC, Hoya M, Wiggins GAR, et al. Using the ACMG/AMP framework to capture evidence related to predicted and observed impact on splicing: Recommendations from the ClinGen SVI Splicing Subgroup. Am J Hum Genet 2023;110(7):1046-1067. DOI: 10.1016/j.ajhg.2023.06.002.
